# Dissecting the Shared Genetic Architecture of Modifiable Physical Activity and Autoimmune Diseases

**DOI:** 10.1101/2025.03.03.25323217

**Authors:** Minjing Chang, Kaixin Yao, Jun Qiao, Yinqi Long, Xicheng Yang, Le Zhou, Qifan Feng, Xi Qiao, Peifeng He, Zhixiu Li

**Affiliations:** School of Public Health and Emergency Management, Southern University of Science and Technology, Shenzhen, China; Department of Nephrology, Shanxi Kidney Disease Institute, Second Hospital of Shanxi Medical University, Taiyuan, China; Department of Pharmacology, School of Medicine, Southern University of Science and Technology, Shenzhen, China; Department of Rheumatology, Second Hospital of Shanxi Medical University, Taiyuan, China; Shanxi Key Laboratory of Big Data for Clinical Decision, Shanxi Medical University, Taiyuan, China; Institute of Medical Data Sciences, Shanxi Medical University, Taiyuan, China

**Keywords:** Physical activities, Autoimmune diseases, Genome-wide association studies, Pleiotropic analysis

## Abstract

**Background:** Autoimmune diseases (ADs) represent a diverse group of chronic conditions that significantly contribute to global disease burden and mortality. Physical activities (PAs) are recognized as key modifiable lifestyle factors for ADs, with numerous epidemiological studies confirming their strong association. Despite this established link, the shared genetic determinants underlying these associations remain largely unexplored.

**Methods:** Using large-scale and comprehensive GWAS summary statistics, we investigated the shared genetic basis between PAs and ADs and explored their vertical and horizontal pleiotropy in detail, including causal relationships, single nucleotide polymorphisms, genes, and biological pathways. Our study revealed extensive genetic associations and overlaps between PAs and ADs.

**Results:** Active PAs generally show negative genetic correlations with ADs, whereas inactive PAs demonstrate positive genetic correlations. From the standpoint of vertical pleiotropy, our results established causal links between specific trait pairs, including Leisure screen time (LST)-Rheumatoid arthritis, LST-Systemic lupus erythematosus, and Sedentary commuting-Type 1 diabetes. In terms of horizontal pleiotropy, we identified 226 loci encompassing 324 significant pleiotropic genes. Notably, GPX1, RHOA, BSN, and MST1 (all located at 3p21.31) emerged as key genes, underscoring the critical roles of T cell activation and lysosomal trafficking pathways in mediating the genetic associations between PAs and ADs.

**Conclusions:** Our findings suggest a shared genetic architecture and underlying mechanisms between PAs and ADs. This study presents the first comprehensive effort of the shared genetic architecture between physical activities and autoimmune diseases but also opens new avenues for the prevention of ADs through increased PAs.

## Background

Autoimmune diseases (ADs) are a heterogeneous group of chronic conditions triggered by an inappropriate immune response to self-antigens, significantly contributing to global morbidity and mortality. Recent studies have highlighted that physical activities (PAs) are a key modifiable risk factor for ADs, and low levels of PAs have a major impact on the burden of disease[1]. PAs are defined as any body movement produced by skeletal muscle that results in energy expenditure and is globally recognized as an important modifiable risk factor and the fourth leading cause of death[2]. For example, A review of 40 studies focusing on systemic lupus erythematosus (SLE) highlighted significant efficacy and tolerance of PAs in stable patients[3]. Moreover, prospective cohort studies have demonstrated an inverse relationship between PA levels and the risk of developing inflammatory bowel disease (IBD) [4]and rheumatoid arthritis (RA)[5]. Lack of PAs can alter the secretion and function of myokines (such as IL-6, irisin, etc.), leading to resistance to their effects and thus inducing a pro-inflammatory response. This state is associated with the occurrence or aggravation of a variety of chronic diseases (such as type 2 diabetes, cardiovascular diseases, ADs, etc.).[6]. Recently, there has been increasing interest in the potential of PAs to maintain immunity and prevent multimorbidity. Although numerous epidemiological studies have established a robust link between PAs and ADs, this association’s precise mechanisms require further exploration.

PAs are a complex polygenic trait, and the heritability estimates of PAs in large twin studies range from 31% to 71%[7]. Using data from 51 studies recently, GWAS identified 99 loci associated with self-reported moderate to vigorous leisure-time physical activity (MVPA), leisure-time screen time (LST), and/or sedentary behavior at work (SDW)[7]. Some of these are shared risk loci with ADs, highlighting the genetic link between PAs and AD phenotypes, so the high association between the two may be attributed to a shared genetic architecture. It is speculated that genetic factors explain their phenotypic associations through mechanisms such as vertical pleiotropy (in which genetic variation affects one trait by affecting another) and horizontal pleiotropy (in which variation affects two traits independently). Previous Mendelian randomization (MR) studies have established causal relationships between specific PAs and ADs. For example, Huang et al. found that reducing long-term leisure TV viewing time has a protective causal effect against (RA)[8]. Conversely, LST has been linked causally to a higher risk of ulcerative colitis (UC)[9]. However, these MR studies primarily focus on vertical pleiotropy and only comprehensively address some traits, thus not fully elucidating the complex genetic relationships between PAs and ADs. Recent developments in genomic statistical methods have highlighted the significance of horizontal pleiotropy in accounting for shared genetic structures beyween human traits. For instance, Gong et al. utilized GWAS databases to explore genetic influences shared along the gut-brain axis, paving the way for identifying common genetic loci associated with lifestyle health aspects (LHA)[10]. Despite these advancements, the shared genetic components between PAs and ADs still need to be explored, with few studies investigating horizontal pleiotropy. Therefore, determining the extent to which genetic factors contribute to the association between PAs and ADs and further exploring their shared genetic architecture and mechanisms are crucial to enhancing our understanding of their mutual relationship. Our findings not only advance theunderstanding of genetic determinants of PAs and ADs but also provide novel insightsinto the shared genetic etiology of PAs and ADs from fuctional and biologicapathway levels.

In this study, we aimed to elucidate the genetic associations and potential causal relationships between PAs and ADs by analyzing the most comprehensive and recent GWAS summary data from individuals of European ancestry[7, 11–15]. First, we uncovered the shared genetic foundation between PAs and ADs using extensive genome-wide genetic correlation, which was further supported by analyses of genetic overlap and regional genetic correlations. We then utilized MR within a vertical pleiotropy framework to assess causality, accounting for confounding effects and sample overlap. Furthermore, we investigated horizontal pleiotropy by detecting pleiotropic variants at the single nucleotide polymorphisms (SNP) level and conducting pairwise colocalization analyses. Subsequent analyses pinpointed candidate pleiotropic genes through positional mapping and eQTL assessments, incorporating chromatin interaction data to enhance gene-based association analyses. Expanding on these results, we conducted pathway enrichment analyses and clarified the biological roles of genes linked to multiple effect loci. In summary, our study establishes a link between PAs and ADs and delves into their shared genetic architecture and biological mechanisms, providing new insights for diagnostic and therapeutic strategies in clinical practice.

## Method

### Study Design

Figure 1 presents the workflow for this study.

**Figure 1:**
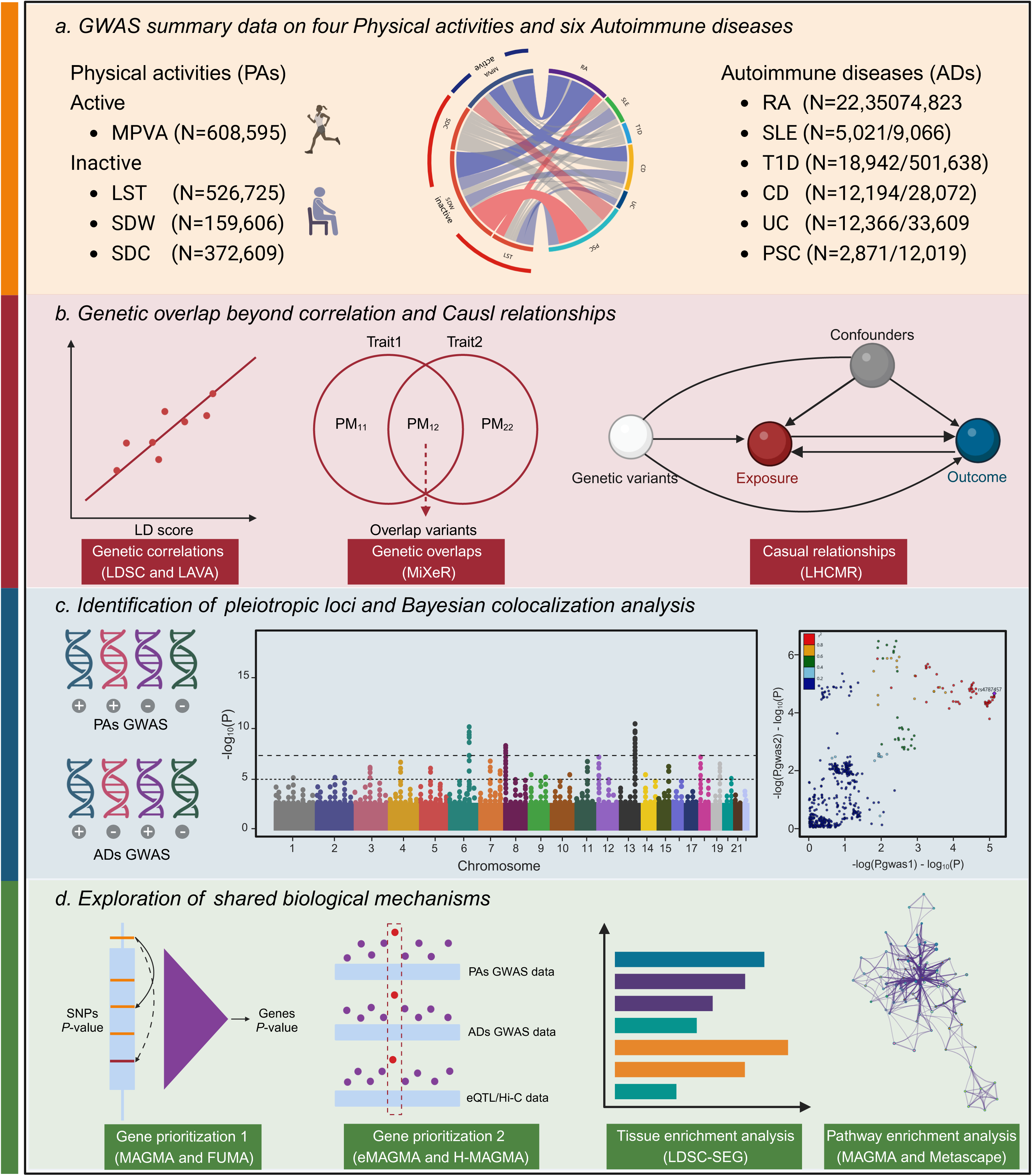
Flowchart of the pleiotropic analysis between four physical activities and six autoimmune diseases. Using large-scale GWAS summary data, we applied a comprehensive genetic approach to examine global and local genetic correlations and overlaps between PAs and ADs, unveiling a shared genetic architecture. To investigate the underlying mechanisms, we analyzed genetic pleiotropy, considering both vertical and horizontal components. For vertical pleiotropy, we conducted an extensive LHCMR analysis to assess causal relationships between PAs and ADs. Additionally, we employed various genetic statistical methods to systematically examine SNPs, genes, and biological pathways, providing insights into horizontal pleiotropy. Our investigation not only identified critical genetic associations but also offered valuable implications for clinical diagnosis and treatment strategies. LST, Leisure screen time; MPVA, Moderate-to-vigorous intensity physical activity during leisure time; SDW, Sedentary behavior at work; SDC, Sedentary commuting; RA, Rheumatoid arthritis; SLE, Systemic lupus erythematosus; T1D, Type 1 diabetes; CD, Crohn’s disease; UC, Ulcerative colitis; PSC, Primary sclerosing cholangitis.

### GWAS datasets for PAs and ADs

The GWAS summary statistics for PAs were derived from an extensive meta-analysis encompassing 51 studies involving up to 661, 399 participants of European ancestry[7]. Specifically, our study included data on LST involving 526, 725 participants, MVPA involving 608, 595 participants, SDW involving 159, 606 participants, and Sedentary commuting (SDC) involving 372, 609 participants. These four PA phenotypes were categorized into two groups: (1) active, which included MVPA, and (2) inactive, which included LST, SDW, and SDC. For ADs, we utilized the latest publicly available GWAS summary statistics for individuals of European ancestry. To ensure higher statistical power and clinical significance, we selected six major ADs. Specifically, the GWAS summary statistics for RA were obtained from a meta-analysis of 37 cohorts, encompassing 22, 350 cases and 74, 823 controls across 25 European cohorts[11]. The summary statistics for SLE were derived from a combination of new GWAS, a meta-analysis of published GWAS, and a replication study involving 5, 201 cases and 9, 066 controls from European subsets[12]. For Type 1 diabetes (T1D), the GWAS summary statistics included 18, 942 cases and 501, 638 controls from a meta-analysis of nine large GWAS[13]. The summary statistics for Crohn’s disease (CD) and UC were sourced from the International IBD Genomics Consortium, including 12, 194 CD cases with 28, 072 controls and 12, 366 UC cases with 33, 609 controls[14]. The GWAS summary statistics for Primary sclerosing cholangitis (PSC) were obtained from the IPSCSG Consortium, comprising 2, 871 cases and 12, 019 controls[15]. Supplementary Table 1 provides more detailed information on the summary statistics for all GWAS. Before further analysis, we performed stringent quality control on our genetic data through several critical steps: (1) aligning the data with the European reference from the 1000 Genomes Project v3 constructed on the hg19 genome; (2) excluding non-biallelic SNPs; (3) removing SNPs that either lacked rsIDs or had duplicate rsIDs; and (4) retaining only SNPs with a minor allele frequency (MAF) greater than 0.01. Following these criteria, we included 4, 522, 260 SNPs for the subsequent analyses (Table 1 and Supplementary Table 1).

**Table 1:** Overview of data sources for four physical activities and six autoimmune diseases included in this study. Note: The table shows an overview of the four physical activities and six autoimmune diseases, the abbreviations used in the manuscript, the f publication, the sample size, population, and reference genome on which the summary statistics were based, and the number of SNPs ed in the original summary statistics before we applied filtering.

| Trait | Abbreviations | Year | Population | N | N case | N control | Reference genome | Original no. of SNPs |
| --- | --- | --- | --- | --- | --- | --- | --- | --- |
| Leisure screen time | LST | 2022 | European | 526,725 | NA | NA | GRCh37 (hg19) | 16,806,192 |
| Moderate-to-vigorous intensity physical activity during leisure time | MPVA | 2022 | European | 608,595 | NA | NA | GRCh37 (hg19) | 16,807,564 |
| Sedentary behavior at work | SDW | 2022 | European | 159,606 | NA | NA | GRCh37 (hg19) | 16,801,707 |
| Sedentary commuting | SDC | 2022 | European | 372,609 | NA | NA | GRCh37 (hg19) | 16,791,178 |
| Rheumatoid arthritis | RA | 2022 | European | 97,173 | 22,350 | 74,823 | GRCh37 (hg19) | 13,297,690 |
| Systemic lupus erythematosus | SLE | 2015 | European | 14,267 | 5,201 | 9,066 | GRCh37 (hg19) | 7,915,251 |
| Type 1 diabetes | T1D | 2021 | European | 520,580 | 18,942 | 501,638 | GRCh37 (hg19) | 62,115,237 |
| Crohn's disease | CD | 2017 | European | 40,266 | 12,194 | 28,072 | GRCh37 (hg19) | 9,570,787 |
| Ulcerative colitis | UC | 2017 | European | 45,975 | 12,366 | 33,609 | GRCh37 (hg19) | 9,588,016 |
| Primary sclerosing cholangitis | PSC | 2016 | European | 14,890 | 2,871 | 12,019 | GRCh37 (hg19) | 7,891,613 |

**Note.** The table shows an overview of the four physical activities and six autoimmune diseases, the abbreviations used in the manuscript, the year of publication, the sample size, population, and reference genome on which the summary statistics were based, and the number of SNPs included in the original summary statistics before we applied filtering.

### Genetic correlation between PAs and ADs

To estimate the shared genetic basis of four PAs and six major ADs, we employed linkage disequilibrium score regression (LDSC)[16] to assess their SNP-based heritability (*h^2^_SNP_*) and genome-wide genetic correlation (*r_g_*). LDSC quantifies genetic associations by regressing the product of the marginal z-scores from the GWAS summary statistics for two traits onto the linkage disequilibrium (LD) score for each SNP. First, we performed univariate LDSC to estimate *h^2^_SNP_*, which reflects the proportion of phenotypic variance in a trait attributed to common genetic variants included in the analysis. We used the 1000 Genomes Project Phase 3 European population as a reference panel, excluding the major histocompatibility complex (MHC) region (chr6:26-34 Mb) to minimize bias. Next, we conducted bivariate LDSC analysis to estimate *r_g_*, defined as the covariance in SNP-heritability across all trait pairs. The regression slope from this analysis provides an unbiased estimate of genetic correlation, even when there is sample overlap across two GWAS datasets. Given the number of trait pairs tested, we set a Bonferroni-corrected significance threshold at *P* < 2.08×10^-3^ (0.05 / 24) to adjust for multiple comparisons.

To evaluate the tissue specificity of PAs and ADs, we used stratified LDSC applied to specifically expressed genes (LDSC-SEG)[17]. LDSC-SEG was used to identify enrichments in tissue-specific gene expression and chromatin modifications, leveraging multi-tissue gene expression data from the Genotype-Tissue Expression (GTEx) project and the Franke lab, and chromatin profiling data from the Roadmap Epigenomics and ENCODE projects, respectively. Specifically, we investigated if regions with high heritability of disease or phenotype within each tissue were enriched for specific expression, indicating a potential association between the tissue and the disease or phenotype. The GTEx project supplied tissue-specific expression statistics for 49 tissues, enhanced by integrating histone mark annotations, including DNase hypersensitivity and chromatin marks like H3K27Ac, H3K4me1, H3K4me3, H3K9ac, and H3K36me3, from the Roadmap Epigenomics and ENCODE projects. Each dataset underwent false discovery rate (FDR) correction to ensure robust results, with a significance threshold set at FDR < 0.05.

### Genetic overlap analysis between PAs and ADs

The genetic correlation measured by LDSC may underestimate the shared genetic underpinnings of PAs and ADs because it does not differentiate between mixtures of concordant and discordant genetic effects and an absence of genetic overlap. We applied the causal mixture modeling approach (MiXeR)[18] analysis to address this limitation and quantify the polygenic overlap using GWAS summary statistics. MiXeR estimates the total number of shared and trait-specific ‘causal’ SNPs based on the distribution of z-scores and detailed modeling of LD structure. First, we conducted univariate MiXeR analyses to determine polygenicity (the number of ‘causal’ variants) and discoverability (the average magnitude of additive genetic associations among these variants). For this analysis, we used the 1000 Genomes Project v3 as our reference panel for European samples, excluding the MHC region (chr6:26-35 Mb) to minimize bias. We then performed bivariate MiXeR analyses to quantify the polygenic overlap between traits. Results were visualized in Venn diagrams, depicting the proportion of shared and unique variants accounting for 90% of SNP heritability in each GWAS. Unlike genetic correlations, MiXeR’s estimates of shared ‘causal’ variants are not influenced by the direction of effects on the trait pairs. The genetic variants are categorized into four groups: shared ‘causal’ variants, unique ‘causal’ variants for each trait, and non-causal variants. Furthermore, we calculated Dice coefficients to estimate the similarity between the genetic architectures of the two phenotypes, ranging from 0 to 1, with higher values indicating greater polygenic overlap. The proportion of SNPs with concordant effects among the shared components was evaluated to provide evidence of the mixture effect. Additionally, MiXeR computed the genome-wide correlations across all SNPs (*r_g_*) and the correlation of effect sizes within the shared genetic component (*r_g_s*). We generated modeled versus actual conditional quantile-quantile (Q-Q) and log-likelihood plots to assess model fit visually. The Akaike information criterion (AIC) was applied to evaluate the predictive accuracy of the MiXeR model compared to actual GWAS data. A negative AIC value indicates inadequate model discrimination, suggesting that the MiXeR model cannot be distinctly differentiated from scenarios of maximum or minimum possible overlap.

### Local genetic correlation between PAs and ADs

When genome-wide associations are assessed using LDSC, which only considers the average of shared signals across the entire genome, local genetic associations (local-*r_g_s*) with opposite directions may cancel each other out. To address this issue, we performed the Local Analysis of [co]Variant Annotation (LAVA)[19] to capture this missing dimension by assessing local-*r_g_s* for PAs and ADs. LAVA is a comprehensive framework that can evaluate local heritability for all traits of interest and test conditional genetic relationships between several traits using partial correlation or multiple regression in addition to standard bivariate local *r_g_s* between two traits. Unlike MiXeR, which estimates the direction of effect for all shared ‘causal’ variants, LAVA complements MiXeR by estimating local-*r_g_s* in 2, 495 local LD blocks, each with an average size of 1 Mb. This approach allows for more efficient identification of mixed effect directions. Initially, we performed LAVA-based univariate analyses to test for univariate local genetic signals (i.e., local-*h^2^*). Using a *p*-value threshold of 1×10^-4^, we filtered out sites lacking sufficient univariate heritability, retaining regions with significant SNP heritability. We then conducted 3, 098 bivariate tests to estimate local-*r_g_s* between PAs and ADs. We followed the LAVA analysis protocol outlined in the original article, using an LD reference panel from the 1000 Genomes Project Phase 3 European ancestry sample while excluding the MHC region (chr6: 25-35 Mb). *P*-values of local genetic correlations were adjusted using the FDR method, with an FDR threshold of < 0.05 denoting statistical significance.

### Estimation of causal effects using LHCMR

To explore the potential causal relationship between PAs and ADs, we employed the Latent Heritable Confounder Mendelian Randomization (LHC-MR)[20] to characterize vertical pleiotropy. LHC-MR utilizes comprehensive genome-wide association data—not just genome-wide significant sites—to assess causality and correct for sample overlap more effectively. LHC-MR builds upon and improves traditional two-sample MR by using structural equation models to connect genome-wide associations with traits and confounders. Thus, LHC-MR can simultaneously estimate unbiased bidirectional causal effects between traits and assess confounding effects. Results from LHC-MR are presented as odds ratios (ORs) with 95% confidence intervals (CIs). We determined causal relationships using Bonferroni correction for multiple testing, setting a threshold at *P* = 0.05 / 24 / 2 = 1.04×10^-3^. A unidirectional causal relationship is indicated when the *P*-value is below this threshold, and the *P*-value in the opposite direction exceeds 0.05. Conversely, a bidirectional causal relationship is established when *P*-values in both directions are simultaneously below 1.04×10^-3^. To validate our findings, we conducted sensitivity analyses using several methods: the inverse variance weighted (IVW) method, weighted median, MR-Egger, simple model, and weighted model. Statistical significance was determined at *P* < 0.05.

### SNP-level association analysis

#### Pairwise pleiotropic analysis using PLACO

To explore the horizontal pleiotropic between PAs and ADs, we used Composite Null Hypothesis Pleiotropy Analysis (PLACO) to identify pleiotropic SNPs associated with these paired traits, elucidating the common genetic mechanisms underlying them. PLACO is a novel statistical method that identifies pleiotropic SNPs between two traits by testing a composite null hypothesis that a SNP is associated with zero or one of the traits[21]. This method requires the product of the Z statistics from two studies as input, which is assumed to follow a mixed distribution[22]. PLACO addresses the problem by splitting the composite null hypothesis into three sub-null hypotheses: H00: The SNP is not associated with either trait; H10: The SNP is associated with the first trait but not the second; H01: The SNP is not associated with the first trait but is associated with the second. The alternative hypothesis (H11) states that the SNP is associated with both traits, indicating the presence of a pleiotropic association[23]. SNPs with *P_PLACO_* < 5 × 10^−8^ in PLACO were considered significant pleiotropic variants.

### Genomic Loci Characterization and Functional Annotation

We used the Functional Mapping (FUMA) and Annotation of Genome-Wide Association Studies platform to identify independent loci associated with PAs and ADs and to annotate all candidate SNPs within these regions[24]. FUMA processes GWAS summary statistics and provides annotation, prioritization, and visualization of the results. First, FUMA identifies independent significant SNPs by applying a genome-wide significance threshold of *P* < 5.0 × 10^−8^ and ensuring LD of r² < 0.6. It further refines this by defining a subset of lead SNPs based on mutual independence (r² < 0.1). To delineate distinct genomic loci, all major SNPs with physical overlap were merged if their LD blocks were less than 500 kb apart. The SNP with the lowest P-value within each locus was selected as the TopSNPs. LD information was derived from the 1000 Genomes Project Phase III data based on European ancestry[25]. The directional effects of these shared loci were assessed by comparing the Z-scores of PAs and ADs traits, and we considered them as novel findings if they were inconsistent with any previously reported PAs and ADs loci in existing GWAS.

To predict the functional outcomes of SNPs, we utilized databases with known functional annotations, including Annotate Variation (ANNOVAR) for general categorization and Combined Annotation Dependent Depletion (CADD) scores to assess the potential deleteriousness of SNPs—considering scores above 12.37 as potentially deleterious[26]. We also incorporated RegulomeDB (RDB) scores, highlighting the regulatory functions of SNPs based on eQTLs and chromatin states, with lower scores indicating higher regulatory potential[27]. Moreover, the ChromHMM tool was used to characterize the regulatory landscape across 127 epigenomes, utilizing 5 chromatin tags to predict 15 classes of regulatory states. Additionally, we implemented two gene mapping strategies to associate SNPs with genes: (i) mapping based on physical proximity within a 10 kb window and (ii) leveraging eQTLs from the GTEx database.

### Bayesian colocalization analysis using COLOC

To determine potential shared causal variants in each pleiotropic locus, we performed Bayesian colocalization analysis on the pleiotropic loci annotated by FUMA. COLOC evaluates the presence of a single causal variant assumption and provides posterior probabilities (PP) for five hypotheses per pleiotropic locus: no causal variant for both traits (PPH0), a causal variant only for one trait (PPH1), a causal variant only for the other trait (PPH2), different causal variants for each trait (PPH3), and a common causal variant (PPH4)[28]. The SNP with the highest PPH4 within each locus was selected as the candidate causal variant. If the PPH4 of the pleiotropic locus was ≥ 0.7, we considered the probability of sharing the same causal variant between the two traits to be high[29].

### Gene association analysis

#### Identication of candidate pleiotropie genes using MAGMA

Leveraging results from PLACO and the original single-trait GWAS, we used Multi-marker Analysis of GenoMic Annotation (MAGMA)[30] to map identified loci to nearby genes. MAGMA employs a multivariate regression model to evaluate the additive effects of individual variant associations, assigning SNPs located within the gene body or in the ±10 kb flanking region to the corresponding genes. The method aggregates SNP-level signals into gene-level associations by incorporating the *p*-value of each SNP into the test statistic for each gene, while considering the LD between SNPs. To enhance the robustness of the analysis, we restricted it to protein-coding genes with at least 10 SNPs. We used a European ancestry population from the 1000 Genomes Project Phase 3 as a reference and analyzed SNP positions in gene annotations, focusing on 17, 636 autosomal protein-coding genes based on the Genome Reference Consortium human build 37 (hg19). We applied a Bonferroni correction based on the number of protein-coding genes and trait pairs, setting the significance threshold at a *P* < 1.18×10^-7^ (0.05 / 17, 636 / 24). Because of the complex LD patterns within the MHC region (chr6: 25 - 35 Mb), genes in this region were excluded from the analysis.

### Investigation of the tissue-specific genes using EMAGMA and TWAS

To identify functional gene associations that may be overlooked by proximity-based SNP assignment in MAGMA, we employed eQTL-informed MAGMA (e-MAGMA)[31]. This approach integrates signals from PLACO analysis into tissue-specific evaluations to identify risk genes associated with PAs and ADs. E-MAGMA uses the MAGMA statistical framework and multiple linear principal component regression models to map SNPs to genes, leveraging tissue-specific eQTL data from GTEx (v8) to transform genome-wide statistics into gene-level insights. To focus on tissue-specific genes and reduce confounding from other tissues, we selected 24 out of the 47 tissues available in GTEx (v8). The selection included 13 brain-related tissues, three gastrointestinal tissues, and EBV-transformed lymphocytes, liver, lung, musculoskeletal, pancreatic, pituitary, spleen, and whole blood. We determined statistical significance tissue-specific genes using a Bonferroni-corrected *p*-value threshold adjusted for the number of genes and the number of trait pairs tested. For example, the significance threshold for musculoskeletal tissue-specific genes was determined to be *P* < 2.64 × 10^-7^ (0.05 / 7, 882 / 24), and for spleen tissue at *P* < 6.88 ×10^-7^ (0.05 / 3, 030 / 24).

We conducted a Transcriptome-wide Association Study (TWAS)[32] to explore the association between gene expression in specific tissues and the risk of PAs and ADs utilizing single-trait GWAS results. TWAS integrates eQTLs with summary association statistics from large-scale GWAS to identify genes that regulate complex traits in cis. We employed advanced FUSION software to estimate potential associations between predicted gene expression, gene prediction model weights, and SNP LD matrices. Our study focused on the 24 tissues identified above, ensuring a comprehensive understanding of the relationship between gene expression and disease risk in multiple key human systems, providing a solid foundation for revealing the molecular mechanisms behind disease occurrence. To ensure the clarity of our findings, genes with duplicate names and non-protein-coding genes were removed from the analysis. The final gene set for each tissue was corrected for multiple testing using the Bonferroni method, defining statistical significance as a *p*-value less than 0.05 divided by the number of gene sets tested.

### Mapping GWAS data to genes using H-MAGMA combined with chromatin interaction data

Most brain-associated risk variants identified by GWAS are within the noncoding genome, posing challenges for elucidating their biological mechanisms. To enhance the accuracy of assigning these genetic variants to their target genes in PAs and ADs, we employed the Hi-C coupled MAGMA (H-MAGMA)[33] method. This approach analyzes gene regulatory relationships within disease-relevant tissues, aiding in identifying genes associated with noncoding variants. H-MAGMA improves upon traditional MAGMA by assigning noncoding SNPs to appropriate genes based on long-range chromatin interactions, utilizing Hi-C profiles from human brain tissues to detail the cell type specificity of these genes. Exonic and promoter SNPs are directly linked to their target genes according to their genomic locations, with promoter regions defined as being 2 kb upstream from the transcription start sites of each gene isoform. We conducted chromatin interaction analyses using H-MAGMA, leveraging six Hi-C datasets from the WonLab GitHub repository, which encompass adult brain tissue, midbrain dopaminergic neurons, cortical neurons, fetal brain tissue, iPSC-derived astrocytes, and iPSC-derived neurons. To ensure rigor in our findings, we applied a stringent Bonferroni correction for multiple testing of protein-coding genes and traits, establishing a threshold of *P* < 1.18×10^-7^ (0.05 divided by 17, 636 and then by 24) for statistical significance.

### Pathway-level analysis using MAGMA and Metascape

We conducted MAGMA gene set analysis[30] to explore the biological significance of pleiotropic genes linked to PAs and ADs. MAGMA conducts a competitive gene set association analysis for each trait, adjusting for gene size, variant density, and LD structure. This methodology assesses whether the combined association of genes within a specific set and the corresponding phenotype exceeds that observed in a randomly selected gene set of the same size. To assess their biological significance, we compared these pleiotropic genes with the gene set from the Molecular Signature Database (MSigDB), selected 7, 744 Gene Ontology (GO) pathways and 186 Kyoto Encyclopedia of Genes and Genomes (KEGG) pathways. We applied a stringent significance threshold of *P* < 2.64 × 10^-6^, calculated by dividing 0.05 by the product of 7, 930 and 24, to manage multiple tests during our pathway and process enrichment analyses of the gene lists.

To better understand the biological basis between PAs and ADs, we performed Metascape analysis[34] on the genes identified by both MAGMA and e-MAGMA. We applied its powerful functional annotation service, which uses high-quality, cutting-edge data to elucidate signaling pathways associated with gene sets. Metascape enables us to delve deeper into gene functions by integrating rich ontology resources from GO biological processes and KEGG pathways. Pathways with *P* < 0.01 were designated as significant.

## Results

### Genome-wide genetic correlation between PAs and ADs

First, univariate LDSC was performed using summary statistics of PAs and ADs to estimate genome-wide *h^2^_snp_*. We found that LST had the highest heritability among PAs (*h^2^_snp_*= 0.076, SE = 0.0025) and SDC had the lowest heritability (*h^2^_snp_*= 0.025, SE = 0.0040). The heritability of the six major ADs ranged from 3.2% to 58.4%, with SLE having the highest heritability (SE = 0.0797) and T1D having the lowest heritability (SE = 0.0039)(Table 2 and Supplementary Table 2a). Subsequently, bivariate analysis was performed to explore the *r_g_* between PAs and ADs. The results showed that 10 of the 24 pairs of traits were genetically correlated, of which 3 pairs were positively correlated and 7 pairs were negatively correlated. After Bonferroni correction (*P* = 0.05 / 24 combination = 2.083×10^-3^), five pairs of traits remained significant. Of note, among the trait pairs of inactive PA-AD, there were genome-wide positive genetic correlations between SDW and PSC (*r_g_* = 0.315, SE = 0.089; *P* = 4.00 × 10^-4^) and LST and RA (*r_g_* = 0.094, SE = 0.030; *P* = 1.80 × 10^-3^). In contrast, SDW and RA showed an opposite genome-wide negative genetic correlation (*r_g_* = -0.262, SE = 0.043; *P* = 1.41 × 10^-9^), which may be inconsistent due to confounding. For the active PA-AD trait pairs, we found that MVPA and RA showed a high genome-wide negative genetic correlation (*r_g_* = -0.215, SE = 0.039; *P* = 2.34×10^-8^), followed by MVPA and CD (*r_g_* = -0.159, SE = 0.034; *P* = 3.99×10^-6^)(**Table 2** and Supplementary Table 2b).

**Table 2:** Genetic correlation and genetic overlap estimations between four physical activities and six autoimmune diseases. Note. Genetic correlation and genetic overlap were estimated using LDSC, MiXeR and LAVA methods, respectively. Genome-wide genetic correlation: The Bonferroni-corrected significance threshold was set at *P* < 2.83×10^-3^ (0.05/24), and finally a joint set of 5 traits with significant genetic correlation was obtained for subsequent analysis. Genetic overlap: **N_PA_, N_AD_** and **N_PA-AD_** = the number of variants that affect PAs, ADs and overlap respectively; **N_PA-AD_(/N_PA-AD_+N_PA_)** represents the proportion of the number of shared causal variants to the total number of PAs and shared causal variants; **N_PA-AD_/(N_PA-AD_+N_AD_)** represents the proportion of shared causal variants to the total number of ADs and shared causal variants; fraction_concordant_within_shared (se) = consistent effect direction within shared components Proportion of influencing variables (standard error). Local correlation: LAVA estimates the number of independent gene loci with significant genetic correlation (positive/negative correlation) after *P* < 0.05 and FDR < 0.05 respectively. Abbreviations: LST, leisure screen time; MPVA, moderate to high intensity physical activity during leisure time; SDW, sedentary behavior at work; SDC, sedentary commuting; RA, rheumatoid arthritis; SLE, systemic lupus erythematosus; T1D, Type 1 diabetes; CD, Crohn’s disease; UC, ulcerative colitis; PSC, primary sclerosing cholangitis.

| Trait pair | Genome-wide genetic correlation |  | Genetic overlap |  |  | Local genetic correlation |  |
| --- | --- | --- | --- | --- | --- | --- | --- |
| | Genetic correlation (SE) | P value for LDSC | $N_{PA-AD}/(N_{PA-AD}+N_{PA})$ | $N_{PA-AD}/(N_{PA-AD}+N_{AD})$ | fraction_concordant t_within_shared (se) | positive correlation/negative correlation (p<0.05) | positive correlation/negative correlation (FDR<0.05) |
| LST-RA | 0.0938 (0.0300) | $1.80 \times 10^{-3}$ | 0.032 | 0.646 | 0.912 (0.048) | 22/10 | 4/1 |
| LST-SLE | -0.0605 (0.0434) | $1.63 \times 10^{-2}$ | 0.004 | 0.998 | 0.006 (0.016) | 18/25 | 3/2 |
| LST-T1D | 0.0241 (0.0252) | $3.39 \times 10^{-2}$ | 0.013 | 0.549 | 0.674 (0.09) | 22/13 | 2/1 |
| LST-CD | 0.0441 (0.0292) | $1.31 \times 10^{-2}$ | 0.020 | 0.519 | 0.708 (0.112) | 21/21 | 3/2 |
| LST-UC | -0.0697 (0.0334) | $3.70 \times 10^{-2}$ | 0.021 | 0.511 | 0.191 (0.083) | 15/19 | 4/3 |
| LST-PSC | -0.1078 (0.0485) | $2.61 \times 10^{-2}$ | 0.008 | 0.997 | 0.016 (0.035) | 9/17 | 2/6 |
| MPVA-RA | -0.2153 (0.0385) | $2.34 \times 10^{-8}$ | 0.051 | 0.952 | 0.033 (0.039) | 6/15 | 0/5 |
| MPVA-SLE | -0.0487 (0.0526) | $3.54 \times 10^{-2}$ | 0.004 | 0.850 | 0.129 (0.174) | 17/13 | 0/3 |
| MPVA-T1D | -0.0586 (0.0347) | $9.11 \times 10^{-2}$ | 0.015 | 0.540 | 0.308 (0.12) | 7/10 | 1/1 |
| MPVA-CD | -0.1586 (0.0344) | $3.99 \times 10^{-6}$ | 0.038 | 0.879 | 0.105 (0.077) | 10/9 | 2/2 |
| MPVA-UC | -0.0565 (0.0488) | $2.47 \times 10^{-2}$ | 0.028 | 0.613 | 0.175 (0.095) | 8/14 | 1/2 |
| MPVA-PSC | 0.1705 (0.0671) | $1.10 \times 10^{-2}$ | 0.009 | 0.996 | 0.998 (0.006) | 19/11 | 3/2 |
| SDW-RA | -0.2615 (0.0432) | $1.41 \times 10^{-9}$ | 0.098 | 0.894 | 0.062 (0.055) | 10/4 | 1/1 |
| SDW-SLE | -0.125 (0.0588) | $3.35 \times 10^{-2}$ | 0.008 | 0.982 | 0.024 (0.051) | 7/6 | 0/1 |
| SDW-T1D | -0.068 (0.0391) | $8.23 \times 10^{-2}$ | 0.033 | 0.610 | 0.257 (0.099) | 3/4 | 1/0 |
| SDW-CD | -0.1191 (0.0453) | $8.50 \times 10^{-3}$ | 0.046 | 0.528 | 0.191 (0.086) | 8/5 | 2/1 |
| SDW-UC | -0.0211 (0.0647) | $7.44 \times 10^{-2}$ | 0.040 | 0.437 | 0.597 (0.095) | 5/5 | 2/1 |
| SDW-PSC | 0.3150 (0.0891) | $4.00 \times 10^{-4}$ | 0.018 | 0.999 | 0.998 (0.006) | 11/8 | 1/0 |
| SDC-RA | 0.0237 (0.0700) | $7.36 \times 10^{-2}$ | 0.030 | 0.269 | 0.701 (0.200) | 9/12 | 0/0 |
| SDC-SLE | 0.0414 (0.0786) | $5.98 \times 10^{-2}$ | 0.008 | 0.905 | 0.855 (0.207) | 6/8 | 2/1 |
| SDC-T1D | 0.0444 (0.0718) | $5.36 \times 10^{-2}$ | 0.030 | 0.537 | 0.714 (0.122) | 4/3 | 1/0 |
| SDC-CD | 0.1057 (0.0591) | $7.37 \times 10^{-2}$ | 0.045 | 0.511 | 0.725 (0.124) | 8/7 | 1/0 |
| SDC-UC | -0.0262 (0.0656) | $6.90 \times 10^{-2}$ | 0.040 | 0.427 | 0.206 (0.103) | 9/3 | 0/0 |
| SDC-PSC | -0.1758 (0.1089) | $1.06 \times 10^{-2}$ | 0.016 | 0.898 | 0.072 (0.107) | 5/6 | 0/2 |

### Genetic Overlap between PAs and ADs

LDSC revealed the overall genetic correlation but failed to capture the differing effect directions among shared genetic variants. Therefore, we utilized MiXeR to investigate the genetic overlap between PAs and ADs and consider mixed effect directions simultaneously. Through univariate MiXeR, we found that PAs integrally demonstrated higher polygenicity than ADs. For PAs, LST possessed the highest polygenicity (N = 10, 923, ‘causal’ variants explaining 90% of LST’s *h^2^_SNP_*, SD = 574), suggesting the presence of a substantial number of causally-related genomic regions in the polygenic structure of the disease. Whereas RA (N = 533, SD = 47) was the disease with the most polygenic traits among ADs, SLE (N = 41, SD = 5) had the least polygenic traits(Supplementary Table 3a). Subsequent bivariate MiXeR analysis identified polygenic overlap between PAs and ADs, with a Dice coefficient (the ratio of shared variants to total variants) ranging from 0.007 to 0.177.(Table 2, Supplementary Fig. 1, Supplementary Table 3b). Of note, SDW and RA had the most genetic overlap (Dice= 0.177), with an estimated total of 477 (SD = 48) variants shared between SDW and RA, representing 10.20% of SDW-influenced variants and 83.33% of RA-influenced variants, while they have the highest negative genetic correlation (*r_g_*=-0.286, SD=0.0242). A similar relationship occurred in MPVA-RA, MPVA-CD, and SDC-UC (MVPA-RA: Dice=0.097, *r_g_*=-0.218; MVPA-CD: Dice=0.072, *r_g_*=-0.166; SDC-UC: Dice=0.078, *r_g_*=-0.096). These results indicated a strong similarity in the shared genetic architectures of these trait pairs and were consistent with their significant LDSC-estimated genetic correlations. Genetic overlap was also observed in the presence of weak *r_g_*. For example, although SDW and UC had the lowest *r_g_* (0.032, SD=0.026), they shared 197 (SD=122) variants (Dice=0.074), accounting for 4.03% of the variance affecting SDW and 43.68% of the variance affecting UC. Genetic overlap but weak rg indicates mixed effect directions, which can be supported by the MiXeR-estimated proportion of shared ‘causal’ variants with concordant effects (0.60, SD=0.09). An analogous mixed effect direction was also evident between SDC and RA (*r_g_*=0.046, the proportion of shared ‘causal’ variants=0.70). Compared to low polygenic disorders like T1D and RA, LST demonstrated high polygenicity, which results in large differences in the number of shared and unique ‘causal’ variants. For instance, LST and RA shared a smaller number of variants (the number of influential variants for the overlap: 344, SD=43), with many more unique-LST variants (10.6K, SD=0.6K) than unique-RA variants (189, SD=59). Simultaneously, LST-RA possessed a high genetic correlation at the genome-wide level (*r_g_*=0.135, SD=0.015) and shared variants (*r_g_s*=0.95, SD=0.06), consistent with their significant positive genetic correlations estimated by LDSC, suggesting the highly similar effects of LST and RA.

### Local genetic correlation between PAs and ADs

As the complement of MiXeR, LAVA was utilized to calculate further the local genetic correlation (local-rgs) of PAs and ADs and reveal the mixed effects. By performing univariate analysis, we initially identified heritable regions for PAs and ADs in 2, 495 regions(Supplementary Table 4). The results suggested that LST possessed the most regions with significant heritability(n=706), whereas SDC(n=170) had the least significant regions among PAs. For ADs, we obtained 2858 regions with PSC accounting for the highest proportion(n=612), and the T1D(n=357) was the lowest. Subsequently, based on the region possessing significant SNP heritability, 3098 bivariate tests were conducted, revealing that all the trait pairs possessed genomic regions with local-rgs when P<0.05(Table 2 and Supplementary Table 5). Among these, LST-CD possessed the most regions with significant local-rgs with 26 regions exhibiting positive correlation, while 21 showed a negative correlation. Despite lacking significant genome-wide rg, some trait pairs still displayed significant local genetic correlated regions. For instance, although no significant genome-wide rg was found for LST and SLE, we found them to share 18 positively correlated / 25 negatively correlated loci. The equal distribution of positive and negative correlation regions may cancel each other out, leading to the absence of genome-wide significance. This further supported the prominent mixed effect directions in shared genetic variants. A similar relationship was found between LST and UC (15 positively correlated and 19 negatively correlated), and so on. In addition, despite showing strong negative rg, SDW-RA was found to share mixed effect directions (10 positively correlated and 4 negatively correlated regions). With the strict threshold of FDR < 0.05, 22 out of 24 trait pairs possessed significant local-rgs except SDC-RA and SDC-UC. Interestingly, 5 genetic regions associated with more than one pair of traits were detected. Interestingly, we detected 5 genetic regions associated with more than one pair of traits. For example, LD block 464 on chromosome 3 was associated with LST-CD, LST-PSC, MVPA-CD, MVPA-UC, MVPA-PSC, SDW-CD, SDW-UC, and SDW-PSC. By focusing on LD block 297 on chromosome 2, associations with LST-RA and MVPA-RA were detected; LD block 926 on chromosome 6 was associated with MVPA-RA and SDC-SLE; LD block 478 on chromosome 3 and LD block 969 on chromosome 6 showed positive local-rgs for both LST-RA and LST-T1D.

Overall, we systematically assessed the genetic correlation and overlap between PAs and ADs using LDSC, MiexR, and LAVA. These analyses underscored a substantial shared genetic foundation between PAs and ADs and showed that active PA-AD trait pairs were more likely to have negative *r_g_*, while inactive PA-AD trait pairs were more likely to have positive *r_g_*.

### Causal effects identified between PAs and ADs

Our analyses offer valuable insights into the genetic foundations of PAs and ADs, and to delve deeper into the nature of these connections, we employed LHC-MR. This method allowed us to assess pairwise causal relationships between PAs and ADs, while accounting for potential biases and confounding factors. Our findings revealed a significant positive causal effect of RA on LST, with an increase in RA expression associated with a 10.4% higher risk of LST (β: 0.099; 95% CI: 1.058 to 1.151). Conversely, SLE demonstrated a significant reverse causal effect on LST, reducing its risk by 6.9% (β: -0.072; 95% CI: 0.904 to 0.959). Additionally, our analysis identified a robust positive causal link between SDC and T1D, with SDC increasing the risk of T1D by 39.97% (OR: 1.400; 95% CI: 1.206 to 1.624) (Table 3, Supplementary Fig. 2, Supplementary Table 6a). Notably, the IVW method further confirmed the causal relationship between SLE and LST (β = -0.011, SE = 0.005, *P* = 0.024)(Supplementary Table 6b), reinforcing the evidence of vertical pleiotropy that mediates the interactions between these traits.

**Table 3.** LHCMR results between four physical activities and six autoimmune diseases. Note. Table shows the bidirectional LHCMR results for PAs and ADs, and provides the P value, OR, and 95% CI effect size for the forward LHCMR (exposure: PAs, outcome: ADs), and the P value, β, and 95% CI effect size for the reverse LHCMR (exposure: ADs, outcome: PAs). Bonferroni correction was used for multiple testing to determine causality, and the threshold was set at P = 0.05 / 24 / 2 = 1.04×10^-3^. When P_Forward_ < 1.04×10^-3^ and P_Reverse_ > 0.05, it indicates a forward causal relationship. Vice versa. When P_Forward_ < 1.04×10^-3^ and P_Reverse_ < 1.04×10^-3^, it indicates a bidirectional causal relationship. LHCMR, Latent Heritable Confounder Mendelian Randomization; OR, odds ratio; CI, confidence interval; β, beta coefficient; se, standard error.

| Trait pairs | Forward |  |  | Reverse |  |  |
| --- | --- | --- | --- | --- | --- | --- |
| | OR | 95% CI | $P_{Forward}$ | Beta | 95% CI | $P_{Reverse}$ |
| LST-RA | 1.113 | 0.995-1.246 | 0.061 | 0.099 | 1.058-1.151 | $4.14 \times 10^{-6}$ |
| LST-SLE | 1.270 | 0.888-1.818 | 0.19 | -0.072 | 0.904-0.959 | $1.80 \times 10^{-6}$ |
| LST-T1D | 1.017 | 0.958-1.079 | 0.581 | 0.087 | 0.982-1.211 | 0.105 |
| LST-CD | 1.154 | 0.969-1.374 | 0.109 | 0.047 | 1.017-1.079 | $1.94 \times 10^{-3}$ |
| LST-UC | 0.834 | 0.695-1.001 | 0.051 | -0.042 | 0.9-1.021 | 0.186 |
| LST-PSC | 0.859 | 0.716-1.030 | 0.102 | -0.041 | 0.885-1.04 | 0.314 |
| MPVA-RA | 0.786 | 0.628-0.983 | 0.035 | -0.044 | 0.914-1.001 | 0.055 |
| MPVA-SLE | 0.642 | 0.191-2.158 | 0.473 | -0.038 | 0.859-1.08 | 0.515 |
| MPVA-T1D | 1.084 | 0.981-1.197 | 0.114 | -0.082 | 0.862-0.985 | 0.017 |
| MPVA-CD | 0.805 | 0.507-1.277 | 0.357 | -0.009 | 0.97-1.013 | 0.433 |
| MPVA-UC | 1.182 | 0.667-2.093 | 0.567 | -0.037 | 0.856-1.085 | 0.545 |
| MPVA-PSC | 1.511 | 1.083-2.107 | 0.015 | 0.062 | 1.028-1.102 | $4.18 \times 10^{-4}$ |
| SDW-RA | 0.679 | 0.520-0.886 | $4.43 \times 10^{-3}$ | -0.06 | 0.906-0.98 | $2.68 \times 10^{-3}$ |
| SDW-SLE | 0.386 | 0.135-1.105 | 0.076 | 0.012 | 0.982-1.043 | 0.429 |
| SDW-T1D | 0.890 | 0.775-1.021 | 0.096 | -0.072 | 0.816-1.061 | 0.281 |
| SDW-CD | 0.544 | 0.162-1.831 | 0.326 | 0.032 | 0.976-1.093 | 0.26 |
| SDW-UC | 1.263 | 0.901-1.772 | 0.176 | 0.055 | 0.905-1.233 | 0.487 |
| SDW-PSC | 0.821 | 0.472-1.427 | 0.484 | 0.055 | 0.96-1.163 | 0.26 |
| SDC-RA | 1.014 | 0.877-1.172 | 0.855 | -0.001 | 0.947-1.055 | 0.983 |
| SDC-SLE | 1.491 | 0.928-2.393 | 0.098 | -0.007 | 0.966-1.021 | 0.61 |
| SDC-T1D | 1.400 | 1.206-1.624 | $9.41 \times 10^{-6}$ | 0.054 | 0.951-1.171 | 0.309 |
| SDC-CD | 0.717 | 0.025-20.323 | 0.846 | 0.029 | 0.946-1.12 | 0.502 |
| SDC-UC | 0.891 | 0.595-1.334 | 0.576 | -0.038 | 0.905-1.024 | 0.225 |
| SDC-PSC | 1.052 | 0.39-2.835 | 0.921 | -0.025 | 0.865-1.099 | 0.676 |

**Note.** Table shows the bidirectional LHCMR results for PAs and ADs, and provides the P value, OR, and 95% CI effect size for the forward LHCMR (exposure: PAs, outcome: ADs), and the P value, $\beta$ , and 95% CI effect size for the reverse LHCMR (exposure: ADs, outcome: PAs). Bonferroni correction was used for multiple testing to determine causality, and the threshold was set at $P = 0.05 / 24 / 2 = 1.04 \times 10^{-3}$ . When $P_{Forward} < 1.04 \times 10^{-3}$ and $P_{Reverse} > 0.05$ , it indicates a forward causal relationship.

### Pleiotropic genomic loci identified between PAs and ADs

We conducted a horizontal pleiotropy analysis using PLACO, identifying 9, 851 significant pleiotropic SNPs (*P* < 5×10^-8^) between four PAs and six ADs. These SNPs were then categorized by FUMA into 262 lead SNPs across 226 loci, spanning 96 distinct chromosomal regions (Fig. 2, Supplementary Fig. 3, Supplementary Table 7). Notably, 16 of these pleiotropic loci have not been previously identified in studies of PAs and ADs, which examined 131 and 43 loci, respectively. This novel identification may stem from variations in the traits or datasets analyzed, differences in analysis methodologies, or definitions of genomic loci, thus providing more robust statistical evidence for pleiotropic associations. The majority of loci, 210 in total, corresponded with regions previously identified as significant in GWAS for at least one of the traits. Among these, the loci at 6p22.1 (*NKAPL*), 3p21.31 (*GPX1*), and 6p21.31 (*MLN*) demonstrated extensive pleiotropic effects, showing overlap in at least half of the trait pairings. In our analysis of 226 pleiotropic loci, we observed that the top-ranked SNPs exhibited mixed directions of allelic association. Specifically, 60 SNPs (27%) increased the risk for both traits, 43 SNPs (19%) decreased the risk, and the remaining 123 SNPs (54%) displayed inconsistent associations. Upon functional annotation using FUMA, we categorized 128 SNPs (48.2%) as intronic variants, 70 SNPs (30.9%) as intergenic, and only 10 SNPs (4.3%) as exonic. Notably, 18 SNPs, encompassing 12 unique genomic risk sites, showed potential harm (CADD score > 12.37), with rs2298428 scoring the highest at 23.9, impacting genes such as *UBE2L3* and *CCDC116*. Additionally, we identified 25 SNPs (representing 8 unique sites) that could affect transcription factor binding, as evidenced by a RegulomeDB score of less than 3; rs4840568 presented the strongest evidence with a score of 1b. Conversely, 42 SNPs had a RegulomeDB score of 7, indicating minimal regulatory potential (Supplementary Fig. 4). Further analysis revealed that 32 of the 226 loci (14.2%) had high posterior probabilities (PPH4 > 0.7) of association with paired traits, indicating significant pleiotropy. Notably, the locus at 3p21.31 was implicated in multiple trait pairs, including SDW-CD, SDW-PSC, and SDW-UC, with PP.H4 values ranging from 0.920 to 0.943 (Supplementary Fig. 5, Supplementary Table 7).

**Figure 2:**
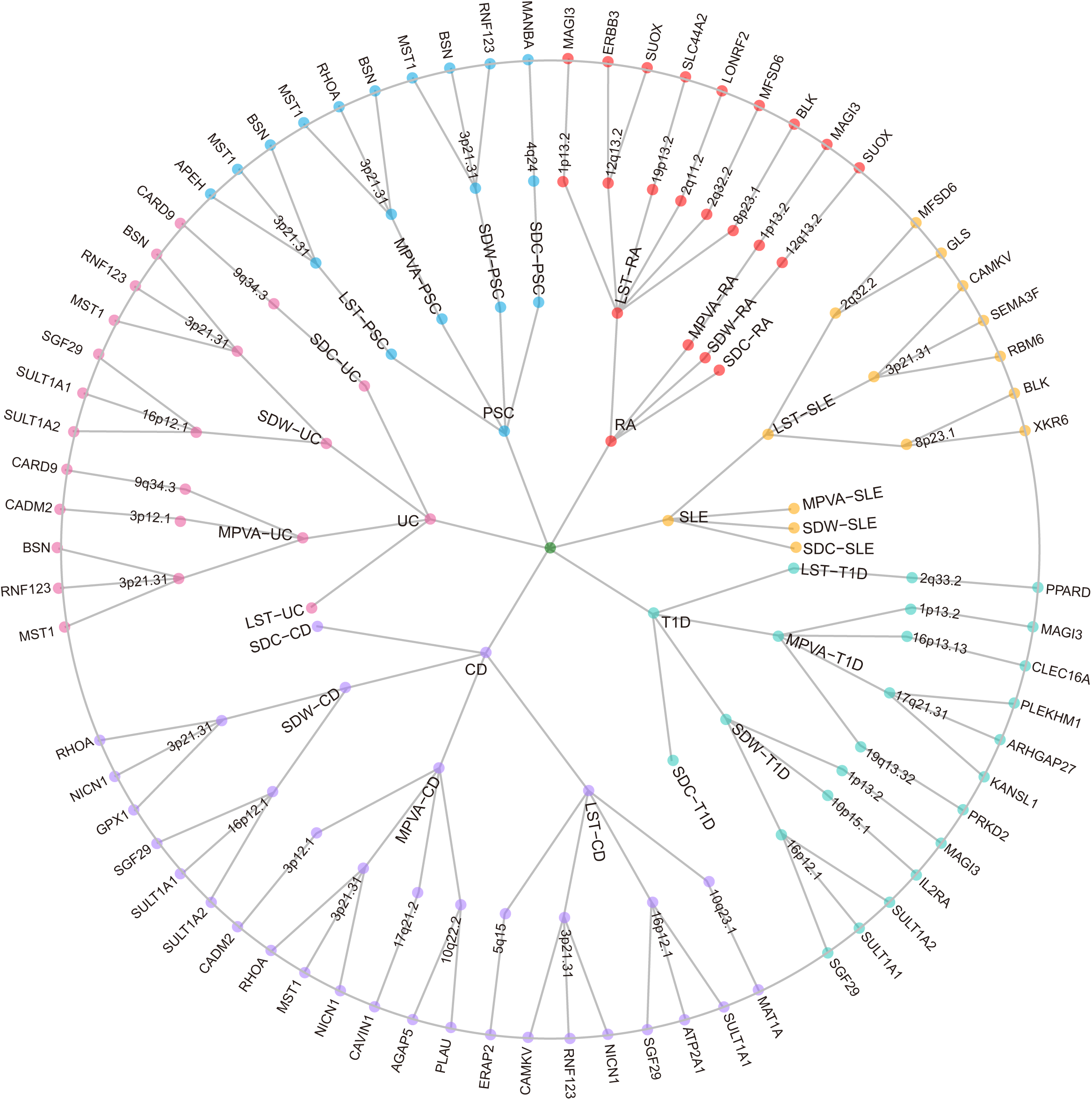
Overview of the pleiotropy associations between four physical activities and six autoimmune diseases. The network diagrams illustrate the identified pleiotropic loci and genes associated with six autoimmune diseases, represented in the inner circle. These loci are connected to four physical activity traits in the second circle, resulting in a total of 24 trait pairs. The third circle showcases 39 pleiotropic loci mapped to 213 significant pleiotropic genes, of which 67 are unique, as determined through the multimarker analysis of genomic annotation (MAGMA). For trait pairs that involve three or more pleiotropic genes, only the top three candidate genes are displayed in the fourth circle, prioritized based on specified criteria. LST, Leisure screen time; MPVA, Moderate-to-vigorous intensity physical activity during leisure time; SDW, Sedentary behavior at work; SDC, Sedentary commuting; RA, Rheumatoid arthritis; SLE, Systemic lupus erythematosus; T1D, Type 1 diabetes; CD, Crohn’s disease; UC, Ulcerative colitis; PSC, Primary sclerosing cholangitis.

### Identification of pleiotropic genes between PAs and ADs

To investigate the mechanisms through which the identified loci may influence PAs and ADs, we employed a MAGMA analysis across 226 loci. This analysis yielded 324 significant pleiotropic genes, including 99 unique genes, using a Bonferroni-corrected significance threshold of *P* = 1.18×10^-7^. Of these genes, 61 are associated with multiple trait pairs, highlighting a common genetic foundation across different conditions(Fig. 2, Supplementary Table 11). A notable concentration of genes was found at the 3p21.31 locus, representing 68.82% of the total identified genes, including *GPX1*, *AMT*, *TCTA*, and others, each associated with at least eight trait pairs. For instance, *TCTA*, which encodes peptide A that inhibits osteoclastogenesis and affects bone remodeling, also impacts the proliferation of lung epithelial cells and synovial fibroblasts in RA[35], potentially reducing inflammation and joint damage. Similarly, *GPX1*, known for encoding an antioxidant enzyme essential in mitigating oxidative stress, has been linked to IBD[36], illustrating its role in both PAs and ADs. Additionally, our study introduces 163 genes previously unexplored in PAs and 45 newly associated with ADs. Using the position mapping feature of FUMA, we further identified 1, 634 potential susceptibility genes, with 97.22% of the MAGMA-identified genes corroborated by the FUMA position map, reinforcing the robustness and reliability of our findings (Supplementary Table 9).

### Characterization of Phenotype and Tissue Specificity

Using the LDSC-SEG approach, we found significant enrichment of gene expression for PAs in 13 specific tissues after applying a stringent threshold (FDR < 0.05). Notably, LST and MVPA showed enrichment across several brain-related tissues, including the anterior cingulate cortex (BA24), cerebral cortex, and frontal cortex (BA9). Conversely, SDW and SDC did not exhibit significant enrichment in any of the selected tissues. In contrast, ADs demonstrated significant enrichment across all six examined tissues. Notably, RA, SLE, and CD showed high correlations with tissues like EBV-transformed B lymphocytes and small intestinal cells, indicating potential pathological changes. Furthermore, CD was also closely associated with the aorta, whole blood, and sun-exposed skin (lower limbs), suggesting its impact on multiple body systems. Furthermore, our multi-tissue chromatin analysis showed that only SDC lacked significant chromatin enrichment in PAs. LST and MVPA showed significant correlation in various tissues, especially in tissues related to the brain. LST was also enriched in tissues related to the anterior ganglion and pancreatic islets, while the enrichment of SDW was limited to tissues from various adult and fetal brains. ADs were primarily significantly enriched in primary cells from peripheral blood, among which CD was significantly enriched in spleen tissue, UC was closely associated with rectal mucosa donors and colon tissue, and T1D was closely associated with fetal thymus, emphasizing their tissue-specific disease mechanisms. Overall, while PAs were predominantly enriched in brain-related tissues, ADs mainly showed enrichment in immune-related tissues. These insights have guided our selection of 24 tissues for further investigation into tissue-specific pleiotropic genes (Supplementary Fig. 6, Supplementary Table 14).

### Phenotypic and tissue-specific characterization of candidate pleiotropic genes

To further investigate the genetic influences on PAs and ADs, we utilized e-MAGMA to map risk variants to genes, focusing on 24 tissues identified for their relevance. Our analysis revealed 800 tissue-specific genes after Bonferroni correction, including 267 unique genes strongly enriched in at least one tissue. Remarkably, *RBM6* was prominent in over half of the trait pairs. Additionally, genes such as *HYAL3*, *GMPPB*, *UBA7*, *MAPKAPK3*, *NAA80*, *RNF123*, *INKA1*, *AMT*, *GNAT1*, *MST1*, *MST1R*, *SEMA3F*, *CACNA2D2*, *IL27*, and *ZGLP1* were significantly distributed across at least ten trait pairs, indicating widespread genetic effects (Supplementary Table 15). Among these, over 44.75% of the genes, including *HYAL3*, were linked to LST-related trait pairs. *HYAL3*, encoding hyaluronidase 3, is crucial for the integrity of the extracellular matrix, essential in muscle, joint, and tissue stability, implicating it in PAs. Furthermore, *HYAL3*’s interaction with hyaluronic acid[37] enhances inflammatory responses, associating it with an elevated risk of SLE, particularly noted by increased hyaluronic acid presence in renal lesions of SLE patients, underscoring its role in the disease’s renal pathogenesis. Using TWAS, we further explored the tissue-specific expression of these genes in original GWAS results and identified 204 novel genes associated with PAs and 129 novel genes associated with ADs (Supplementary Table 16). eQTL mapping using FUMA revealed 1, 755 genes, 70.75% associated with PAs and ADs, highlighting their pleiotropic roles in disease susceptibility. These findings highlight significant genetic interactions between PAs and ADs while identifying genes with potential tissue-specific effects, providing new insights into these complex traits’ molecular mechanisms (Supplementary Table 9).

In all, our study identified 213 pleiotropic genes (67 unique) at the intersection of MAGMA and e-MAGMA analyses, with over 69.48% of these overlapping genes located at the genetic locus 3p21.3 (Fig.2 and Supplementary Table 11). This region houses critical genes like *GPX1*, *AMT, NICN1*, *APEH*, *MST1*, *RNF123*, *RHOA*, and *BSN*, each implicated in at least eight different trait pairs. Notably, *GPX1* plays a pivotal role in PAs and immune function by maintaining cellular redox balance, mitigating exercise-induced oxidative stress, and modulating inflammatory responses. Similarly, *RHOA* is crucial for motor function[38] as it regulates actin fibers in muscle cells through the activation of Rock and enhances glucose uptake in skeletal muscles via GLUT4 translocation, processes that are potentially linked to ADs. Additionally, the *BSN* gene impacts energy expenditure and storage[39], influencing BMI levels and potentially altering the susceptibility and severity of CD and UC by modulating immune cell function and inflammatory responses[39].

### Mapping GWAS data to genes using H-MAGMA combined with chromatin interaction data

Using H-MAGMA method for chromatin interaction profiling, we identified specific gene enrichments across several tissue cell types: 499 adult brain genes, 396 cortical neuronal genes, 481 fetal brain genes, 382 iPSC-derived astral genes, 365 iPSC-derived neural genes, and 464 DA-midbrain tissue cell type-specific genes. Notably, within these cell types, LST showed substantial enrichment with 1, 051 genes (77.2%) identified, whereas only one gene in SDC was enriched in midbrain DA tissue. In ADs, the most significant enrichments were observed for CD and T1D with 1, 236 (31.8%) and 931 (23.9%) genes respectively, across the six analyzed tissue cell types (Supplementary Table 17). Further, we analyzed the Hi-C interactions between trait pairs and identified 2, 587 significant interactions. Of these, the LST-CD trait pair accounted for the highest number, with 308 interactions (11.9%) (Supplementary Table 18). Additionally, when overlapping the results from MAGMA and H-MAGMA analyses, and restricting to genes at GWAS loci, we identified 1, 195 genes, including 266 unique genes. Among these, seven genes (*GPX1*, *TCTA*, *AMT*, *DAG1*, *BSN*, *MST1*, *CAMKV*, *RHOA*) were notably enriched in six tissues and influenced multiple traits. For instance, variations in *AMT* (aminomethyltransferase) activity can significantly impact ammonia levels[40], thereby influencing immune cell function and inflammatory responses. Abnormal expression of the *AMT* gene disrupts this balance, potentially causing elevated ammonia levels, which can activate immune cells, exacerbate inflammation, and accelerate the progression of ADs.

### Shared Biological mechanism Between PAs and ADs

In our study, we used MAGMA gene set analysis to evaluate pathways of pleiotropic genes associated with PAs and ADs. Using a Bonferroni-corrected threshold of *P* < 5.43×10^-8^, we identified a total of 83 pathways, including 80 GO terms and 3 KEGG pathways. Notably, 17 of these pathways were shared among multiple trait pairs, especially with significant representation in MVPA-RA and MVPA-T1D trait pairs, accounting for nearly half of all identified pathways. Over half of these pathways were closely associated with immunoregulatory processes, such as T cell activation, lymphocyte activation, general immune response, and positive regulation of immune system processes (Supplementary Table 19a). Moreover, we performed functional enrichment analysis using Metascape on the 67 pleiotropic genes that overlapped in the MAGMA and e-MAGMA results to explore their key biological functions. Metascape identified 15 significantly enriched pathways, highlighting immune-related processes, as well as pathways involved in lysosomal trafficking, proteolysis regulation, endocytosis, and macroautophagy. Among them, the lysosomal pathway contains a large number of immune-related genes (such as *MST1*, *RHOA*, *BSN*, *CLN3*, *NUPR1*, and *ATP2A1*), which may be involved in the formation, functional maintenance, and related metabolic processes of lysosomes. This pathway can affect PAs through energy metabolism regulation and affect ADs[41] through its role in antigen processing and presentation (Supplementary Table 19b).

## Discussion

This study conducted a comprehensive genome-wide intersectional trait analysis of four PAs and six major ADs using large-scale GWAS summary statistics. Our systematic investigation uncovered extensive genetic associations and overlaps between PAs and ADs, highlighting potential genetic links often overlooked in phenotype-specific studies. Notably, there is typically a negative genome-wide *r_g_* between active PAs and ADs, in contrast to a positive *r_g_* observed between inactive PAs and ADs. We also pinpointed important pleiotropic loci and genes, such as *GPX1*, *RHOA*, *BSN*, and *MST1*, all situated at 3p21.31, which regulate biological pathways critical to the pathogenesis of both PAs and ADs, such as T cell activation and lysosomal trafficking. Moreover, our MR analysis confirmed causal relationships between specific trait pairs: LST-RA, LST-SLE, SDC-T1D, and SDW-PSC, further confirming the positive causal relationship between inactive PAs and ADs. In conclusion, active PAs may mitigate the occurrence of ADs by inhibiting the anti-inflammatory responses induced by these pathways, whereas inactive PAs tend to increase susceptibility to ADs. This study not only enhances our understanding of the genetic architecture shared between PAs and ADs but also lays a solid foundation for developing preventive strategies, such as increased exercise.

Although numerous studies have shown a widespread association between PAs and ADs, the genetic foundations of this relationship remain unexplored, emphasizing the need for an in-depth examination of their shared genetic architecture. Various analytical approaches based on different model assumptions enable a comprehensive exploration of potential pleiotropic associations from multiple perspectives. LDSC results indicated that 21% of the cross-traits exhibited significant genetic associations, corroborating previously observed genetic contributions. These analyses underscored a substantial common genetic foundation between PAs and ADs, with active conditions tending to display negative *r_g_*, and inactive ones positive *r_g_*. Furthermore, MiXeR analysis confirmed extensive genetic overlap across all phenotypic clusters, despite variability in the significance of genome-wide rg, likely attributable to mixed effect directions. For example, LAVA analysis of LST-CD revealed 26 positively and 21 negatively correlated regions, indicating that although mixed effects might mask clear genetic correlations, the identification of numerous associated loci still supports a genetic link between these trait pairs. In conclusion, our study elucidates a broad common genetic basis between PAs and ADs, providing further evidence of their genetic interconnection.

The common genetic basis between PAs and ADs can be attributed to horizontal pleiotropy and vertical pleiotropy. We first evaluated vertical pleiotropy by exploring the causal relationship between PAs and ADs. Our analysis revealed a significant positive causal relationship between RA and LST. This relationship may exist because patients with ADs often engage in less active PAs due to increased disease activity, joint pain, or fatigue. Conversely, we observed a reverse causal relationship between SLE and LST, a controversial finding that should be interpreted with caution. Contrary to previous research, our results do not establish a causal link between inactive PAs and RA[42]. This discrepancy may stem from differences in data sources, sample overlap, or the limitations of genetic tools used in earlier studies to screen for PAs. Additionally, our findings align with recent genetic analyses in European populations, which indicate no causal relationship between active PAs and SLE[43]. Moreover, we also found that increased SDC results in a higher risk of T1D. This association could be due to prolonged sitting disrupting lipid metabolism, reducing lipoprotein lipase activity, and consequently heightening the likelihood of impaired metabolic activity, leading to increased systemic chronic inflammation[44, 45]. Our study suggests that causal relationships between PAs and ADs are partially influenced by genetic overlap, and the complexity of these relationships differing across specific condition. In conclusion, while inactive PAs may exacerbate ADs by aggravating systemic chronic inflammation, patients with ADs are likely to engage in fewer active PAs due to symptoms like increased disease activity, joint pain, or fatigue.

Given the significant genetic associations, we further investigated how genetic variants influence PAs and ADs through horizontal pleiotropy. We identified 226 genome-wide significant pleiotropic loci; notably, loci such as 3p21.31 (*SEMA3F*) and 6p21.31 (*BAK1*) were evident in half of the trait pairs, underscoring their extensive pleiotropy. *SEMA3F*, a secreted member of the semaphorin family located at 3p21.31, plays a critical role in the pathogenesis of musculoskeletal diseases[46].*HOXA5*, which regulates *SEMA3F* transcription, is frequently downregulated in the synovial tissue of patients with RA, potentially intensifying the aggressive phenotype of RA fibroblast-like synoviocytes[47]. Intriguingly, increases in *HOXA5* expression following fat loss suggest that a reduction in body mass index (BMI) could enhance *HOXA5* levels, thus potentially impacting PAs. Additionally, *BAK1*, situated at 6p21.31 and part of the pro-apoptotic bcl-2 family, is essential for regulating the intrinsic apoptotic pathway and maintaining lymphocyte numbers[48, 49]. These findings of extensive pleiotropy provide strong evidence of genetic interactions between PAs and ADs, setting the stage for more detailed exploration of their genetic underpinnings.

We further mapped SNP annotations to genes and identified key pleiotropic genes that overlapped in MAGMA and EMAGMA analyses, including *GPX1*, *RHOA*, *MST1*, and *BSN* (all located at 3p21.31), which were present in eight trait pairs. Glutathione peroxidase 1 (*GPX1*), a crucial member of the antioxidant enzyme family, helps maintain cellular redox balance by oxidizing glutathione and reducing hydrogen peroxide[50]. Notably, elevated *GPX1* enzyme activity in obese individuals, relative to those with a normal body mass index, suggests an association with lifestyle factors such as sedentary behavior[51, 52]. Additionally, studies indicate that PAs enhance *RHOA* kinase activity in skeletal muscle, reflecting its responsiveness to behavioral changes[38, 53]. Furthermore, Bassoon (*BSN*) and Macrophage Stimulator-1 (*MST1*) are implicated in the susceptibility to CD and UC, respectively[54, 55]. Recent research also suggests that *MST1* may mediate the adverse effects of sedentary behaviors on CAD, highlighting its potential link with PAs[56]. The identification of these pleiotropic loci underscores significant genetic interactions between PAs and ADs, setting the stage for further investigation into their shared genetic mechanisms.

Many common genetic foundations also determine common biological pathways, among which the regulation of T cell activation and lysosomal trafficking pathways is one of the most important biological pathways and is crucial to the impact of PAs and ADs. Key genes such as *RHOA* are key in regulating the T cell activation pathway. Human and animal studies have shown that PAs promote the shift of the Th1/Th2 balance toward Th2 cell dominance, affecting susceptibility to infection, allergy, and autoimmunity[57, 58]. Although Th1 cells are associated with forming various ADs, Th2 cells are associated with specific types of ADs. At the same time, the *RHOA* protein is essential for activating adaptive immune cells (especially pathogen-specific T cells and B cells), which are essential for adaptive pathogen-specific immune responses[38]. In the lysosomal trafficking pathway, genes such as *MST1*, *RHOA*, and *BSN* are necessary to balance autophagy and immune response. Elevated lysosomal enzyme activity is a characteristic of various ADs, and specific lysosomal proteases play a vital role in the lysosomal breakdown of organelles and in reducing and diagnosing RA. The presence of proteases such as B, D, G, K, L, and S in serum and synovial fluid has been proposed as potential diagnostic markers for RA[59–62]. In addition, autophagy is a highly conserved cellular degradation pathway that involves the lysosomal breakdown of organelles and the recycling of macromolecules. Modulating autophagy by PAs can exacerbate or alleviate diseases characterized by abnormal autophagic activity. Studies have shown that exercise can normalize autophagic function, regulate autophagic levels, maintain cellular homeostasis, reduce the production of reactive oxygen species, and alleviate inflammatory responses[63, 64].

Our study also has several limitations. First, we restricted our analysis to individuals of European ancestry to minimize confounding by population structure and admixture. Therefore, the genetic architecture of PAs and ADs in other populations remains largely unexplored. Future studies aim to replicate these findings in more diverse populations. Second, including subjects with both PAs and ADs may influence our assessment of the genetic overlap between these conditions, potentially skewing the interpretation of genetic analyses. Furthermore, our analysis of the pooled GWAS data covers the six major ADs, which do not fully represent their genetic risk structure but still account for a significant portion. Finally, while we have identified several pleiotropic loci and genes, further investigation is required to elucidate their roles in disease pathophysiology.

Our findings demonstrate that lower levels of PAs are associated with a higher risk of developing ADs, and both physical inactivity and sedentary behavior negatively affect the health outcomes of these patients. For instance, patients with RA who exercise regularly experience slower disease progression, improved cardiovascular health, and enhanced joint mobility[65]. Similarly, SLE patients with higher PA levels report improved quality of life and cardiovascular health[66], while regular physical activity in T1D patients reduces the risk of autonomic neuropathy and CVDs[67, 68]. Given the interrelationship between PAs and ADs, improving disease outcomes through PAs may also promote better compliance and a greater inclination for physical exercise. In summary, exercise is crucial for enhancing immune health, optimizing immune cell function, improving antimicrobial activity, and reducing systemic inflammation. Thus, encouraging ADs patients to “sit less and move more” according to their individual needs, alongside drug therapy and clinical care, can substantially improve their quality of life, cardiopulmonary function, and muscle strength while alleviating symptoms such as pain and depression.

## Conclusions

In summary, our comprehensive assessment of PAs and ADs revealed extensive polygenic overlap and a complex common genetic basis. Exploring horizontal pleiotropy, we identified significant pleiotropic loci and genes, including *GPX1*, *RHOA*, *BSN,* and *MST1*—all situated at 3p21.31. Simultaneously, we pinpointed key pathways and immunological mechanisms involved, notably T-cell activation and lysosomal trafficking. Regarding vertical pleiotropy, we corroborated findings from previous MR studies and discovered new causal relationships between trait pairs. These novel insights into the interplay between PA and AD not only advance our fundamental understanding of their shared etiology but also offer promising new avenues for both preventative strategies and the development of targeted therapeutics, potentially leading to the identification of novel targets for intervention and the development of combined lifestyle and pharmacological strategies to combat AD.

## Data Availability

All data produced in the present study are available upon reasonable request to the authors
All data produced in the present work are contained in the manuscript
All data produced are available online at

https://www.ebi.ac.uk/gwas/

## Acknowledgements

Not applicable.

## Abbreviations

ADs: Autoimmune diseases
PAs: Physical activities
LST: Leisure screen time
MVPA: vigorous leisure-time physical activity
SDW: sedentary behavior at work
SDC: Sedentary commuting
SLE: systemic lupus erythematosus
IBD: inflammatory bowel disease
RA: rheumatoid arthritis
UC: ulcerative colitis
T1D: Type 1 diabetes
CD: Crohn’s disease
CD: Primary sclerosing cholangitis
MR: Mendelian randomization
eQTL: expression quantitative trait loci
MAF: minor allele frequency
LDSC: linkage disequilibrium score regression
LD: linkage disequilibrium
MHC: major histocompatibility complex
LDSC-SEG: LDSC applied to specifically expressed genes
FDR: false discovery rate
GTEx: Genotype-Tissue Expression
MiXeR: causal mixture modeling approach
LAVA: Local Analysis of [co]Variant Annotation
LHC-MR: Latent Heritable Confounder Mendelian Randomization
IVW: inverse variance weighted
PLACO: Composite Null Hypothesis Pleiotropy Analysis
FUMA: Functional Mapping
ANNOVAR: Annotate Variation
CADD: Combined Annotation Dependent Depletion
MAGMA: Multi-marker Analysis of GenoMic Annotation
e-MAGMA: eQTL-informed MAGMA
TWAS: Transcriptome-wide Association Study
H-MAGMA: Hi-C coupled MAGMA
MSigDB: Molecular Signature Database
GO: Gene Ontology
KEGG: Genes and Genomes
BMI: body mass index
RDB: RegulomeDB
PP: posterior probabilities
LHA: lifestyle health aspects
SNP: single nucleotide polymorphisms

## Authors’ contributions

M.C., P. H., X.Q. and Z.L. conceptualized and supervised this project and wrote the manuscript. M.C. and K.Y. performed the main analyses and wrote the manuscript. J.Q., Y.L., X.Y., L.Z. and Q.F. performed the statistical analysis and assisted with interpreting the results. X.Q., P.H. and Z.L. provided expertise in Immunology and GWAS summary statistics. All authors provided intellectual content and approved the final version of the manuscript.

## Availability of data and materials

The study used only openly available GWAS summary statistics on physical activities and six autoimmune diseases that have originally been conducted using human data. GWAS summary statistics on PAs (covering LST, MVPA, SDW and SDC) are available GWAS Catalog (GCST90104339, GCST90104341, GCST90104343, and GCST90104345). GWAS summary statistics on ADs (covering RA, SLE, T1D, CD, UC and PSC) are available at the GWAS Catalog (GCST90132223, GCST003156, GCST90014023, GCST004132, GCST004133, and GCST004030).

## Declarations

All authors disclosed no conflict of interest.

## Footnotes

Springer Nature remains neutral with regard to jurisdictional claims in published maps and institutional affiliations.

